# Cryoballoon Ablation of RooF line combined with pulmonary vein Isolation for persistent atrial fibrillation (The CARFI-PerAF Randomized Clinical Trial)

**DOI:** 10.1101/2023.09.06.23295158

**Authors:** Songqun Huang, Yao Zhao, Ruihan Ju, Chao Liu, Shaohua Dong, Aihong Qin, Jiang Cao, Manli Yu, Zhifu Guo, Xinmiao Huang

## Abstract

**Background:** The limited effectiveness of pulmonary vein isolation (PVI) alone using cryoballoon ablation (CBA) led to addictive ablation in procedures of persistent atrial fibrillation (AF) ablation. Roof line (RL) ablation in addition to PVI hold great promise for reduction of AF recurrence after CBA. The randomized controlled CARFI-PerAF trial aimed to prospectively investigate the efficacy of a novel CBA strategy for block of RL and reduction of AF recurrence.

**Methods:** One hundred and ten patients who were diagnosed with persistent AF were randomized into PVI group and PVI+RL group. ‘Quarter balloon ablation technique’ and ‘roof distortion technique’ were used to improve quality of RL ablation. Conduction block of RL was confirmed by both voltage mapping and upper right atrial septum pacing. Primary effectiveness was freedom from AF or atrial tachycardia absent class I/III antiarrhythmic drugs through 12-month follow-up according to ECGs collected by portable device and 24-hour Holter.

**Results:** There was no significant difference in AF recurrence between PVI group and PVI+RL group (63.5% vs 76.2%, P = 0.296) after 532.7 ± 171.0 days of follow-up. However, blocked RL was associated with a significant reduction in risk of AF recurrence in the PVI+RL group (84.0% vs 45.5%, P = 0.025). The shape of RL was the only factor affecting the success rate of RL block. Patients with ‘Regular’ shape of RL predicted a higher rate of RL block than other types (89.7% vs 56.3%, P = 0.014).

**Conclusions:** Blocked roof line ablation was associated with a significant reduction in risk of atrial fibrillation recurrence after cryoballoon ablation. Patients with ‘Regular’ shape of roof line may benefit more from roof line ablation.

## 1 Introduction

Cryoballoon ablation (CBA) has been widely used for pulmonary vein isolation (PVI) that is the cornerstone in persistent atrial fibrillation (AF) ablation^1-4^. However, the limited effectiveness of PVI alone for persistent AF ablation led to addictive ablation to reduce AF recurrence^5^. Roof line (RL) ablation in addition to PVI hold great promise for intervention of atrial electrophysiological and structural substrates^6-8^.

Retrospective and nonrandomized controlled studies found that RL ablation combined with PVI reduce AF recurrence significantly with a premise that the RL should be completely blocked^9, 10^. The risk of atrial flutter after ablation was increased due to unblocked RL. However, complete block of RL is still challenging because of non-visualization of cryoballoon in most 3D-mapping systems^11, 12^.

The randomized controlled CARFI-PerAF trial aimed to prospectively investigate the efficacy of the novel CBA strategy for reduction of AF recurrence. ‘Quarter balloon ablation technique’ and ‘roof distortion technique’ were used in RL ablation for block of RL.

## 2 Methods

The CARFI-PerAF trial (Cryoballoon Ablation of RooF line combined with pulmonary vein Isolation for Persistent Atrial Fibrillation) is a prospective, randomized controlled study to investigate the efficacy of a novel CBA strategy for block of RL and reduction of AF recurrence (URL: https://www.chictr.org.cn/; Unique identifier: ChiCTR2000039746).

### 2.1 Study participants

One hundred and ten patients who were diagnosed with persistent AF, defined as continuous AF documented on electrocardiography or Holter, were prospectively randomized 1:1 into PVI group and PVI+RL group between November 2020 and August 2022. Eligible patients were between 18 and 80 years of age with no thrombus in the left atrial appendage and left atrium. There was no limitation on left atrial diameter and duration of AF. This study was conducted in accordance with the Declaration of Helsinki and was approved by the institutional review board of Shanghai Changhai Hospital (CHEC2020-116). Informed consent was obtained from all participants.

### 2.2 Cryoballon ablation procedure for PVI

Procedures were performed under conscious sedation. The EnSite Precision™ Cardiac Mapping System (Abbott Medical, Inc., St Paul, MN, USA) was used for electroanatomic activation mapping. A steerable 15 Fr sheath (Arctic Front Advance™, Medtronic Inc., Mounds View, MN, USA) was placed in the left atrium (LA) through a single transseptal puncture. A 28 mm second-generation cryoballoon (Medtronic Inc., Mounds View, MN, USA) was advanced into the LA through the sheath with the Achieve™ (Medtronic Inc., Mounds View, MN, USA) catheter as a guiding tool.

Occlusion of the pulmonary veins (PV) was verified by the injection of contrast agent. A bonus freeze of 180 seconds duration was applied if PV potentials were abolished or dissociated from atrial activity in the first 60 seconds after freeze application. In cases of unsuccessful isolation of PV in the first 60 seconds, delivery of the freeze energy was suspended. The CBA was then performed after adjustment of catheters. Additional ablation at the PV antrum could be applied according to anatomy of LA and PVs. Continuous phrenic nerve pacing was performed ten seconds after freeze application at the right PVs. Heparin was applied to maintain the activated clotting time >300 seconds during the entire procedure. Electrical cardioversion was used under propofol anesthesia if AF was not converted to sinus rhythm after PVI.

### 2.3 Cryoballoon ablation procedure for RL block

Angiogram through the steerable sheath showed the profile of the LA roof, the contour of which was depicted on the screen of DSA. Extraction from the steerable sheath should be prohibited to avoided air embolism. With Achieve catheter in the left superior PV, the sheath was rotated clockwise until the cryoballoon was raised beyond the roof contour for contact of the cryoballoon to the LA roof (the ‘roof distortion technique’, distortion of the LA roof implied contact between cryoballoon and LA roof and lesions should be achieved). The sheath and cryoballoon were then advanced toward the left superior PV to cover the lesions made by PVI. A bonus freeze of 120 seconds duration was applied after the upper and anterior contour of the cryoballoon was depicted on the screen of DSA (the ‘Quarter balloon ablation technique’, only the upper and anterior part of the cryoballoon made contact with the LA roof and achieved lesions). The sheath was then further rotated clockwise and ablation was delivered to expand the lesions through LA roof to the right superior PV until rotation limit was reached. The Achieve catheter was then placed in the right superior PV to conduct the cryoballoon to ablate the right part of RL. The upper and anterior contour of the cryoballoon was traced and depicted until the roof line was completely covered by the trajectory. CBA procedure for RL block is shown in Figure 1.

**Figure 1.**
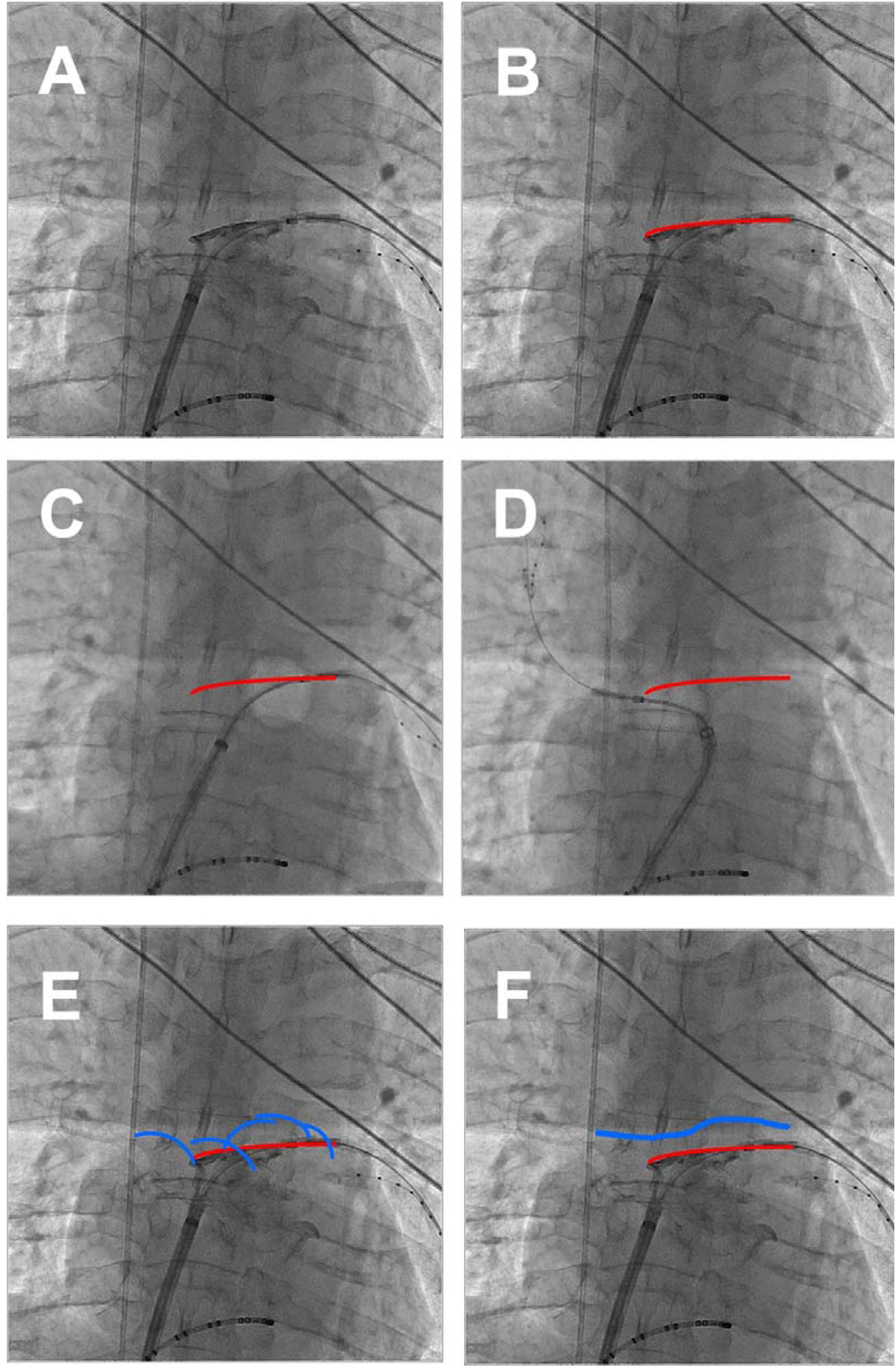
Cryoballoon ablation procedure for roof line block in an example of a representative case. A: Angiogram through the steerable sheath showed the profile of the LA roof in anteroposterior position; B: The contour of LA roof was depicted with red line; C: The ‘roof distortion technique’, the cryoballoon was raised beyond the roof contour for contact of the cryoballoon to the LA roof when the cryoballoon in the left part of the roof line; D: The ‘roof distortion technique’ when the cryoballoon in the right part of the roof line; E: The ‘Quarter balloon ablation technique’, only the upper and anterior part of the cryoballoon made contact with the LA roof and achieved lesions. The upper and anterior contour of the cryoballoon was depicted with blue quarter circle and traced until the roof line was completely covered by the trajectory; F: The trajectory of quarter balloon was connected by blue line that implied the roof distortion during ablation.

### 2.4 Assessment of RL block

Conduction block of RL was assessed by two criteria, both of which should be both fulfilled. First, voltage mapping of LA roof with Achieve catheter showed continuous low voltage (<0.5mv) roof line connecting right and left superior PVs. Second, activation mapping of posterior LA wall with Achieve catheter showed an ascending activation sequence when pacing at the upper right atrial septum with a conduction time >120ms (Figure 2). The conduction time and sequence were also mapped during sinus rhythm. The pacing position of the upper right atrial septum should be adjusted until the difference of conduction time between sinus and pacing rhythm was less than 20ms, implying that the pacing position was on the conduction path of Bachmann’s bundle (Figure 3). If the conduction block of RL was not achieved, the connection positions of the RL revealed by mapping was localized by Achieve catheter on DSA screen and CBA was then applied at that position. If RL block was still not achieved after three attempts of additional ablation, failure of RL ablation was declared. Factors affecting RL block was investigated and RL shaped was classified.

**Figure 2.**
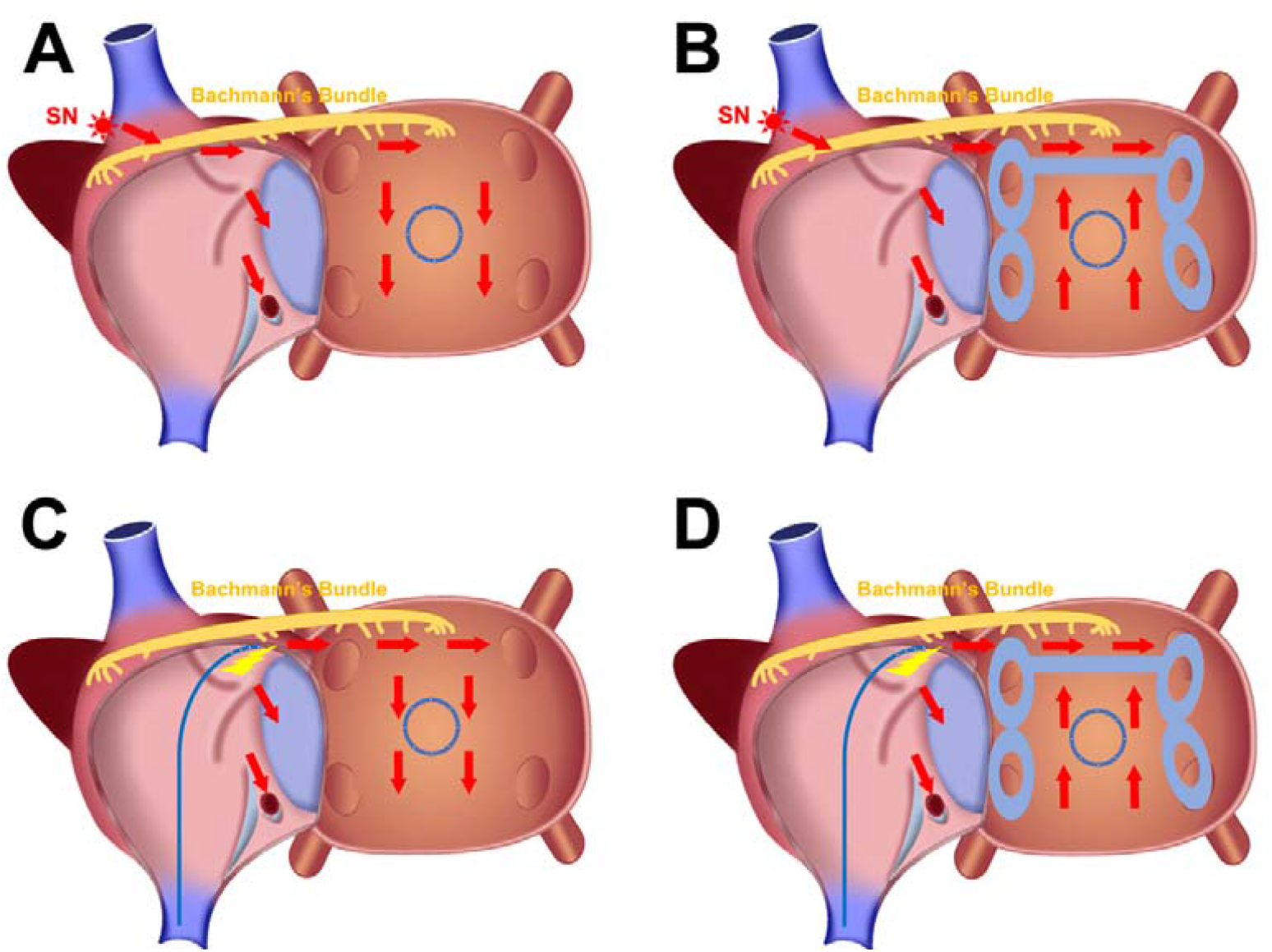
Schematic of mapping from posterior wall of left atrium. A: In sinus rhythm, mapping of posterior wall of LA showed an descending activation sequence before ablation; B: In sinus rhythm, mapping of posterior wall of LA showed an ascending activation sequence after ablation; C: In pacing rhythm, mapping of posterior wall of LA showed an descending activation sequence before ablation; D: In pacing rhythm, mapping of posterior wall of LA showed an ascending activation sequence after ablation

**Figure 3.**
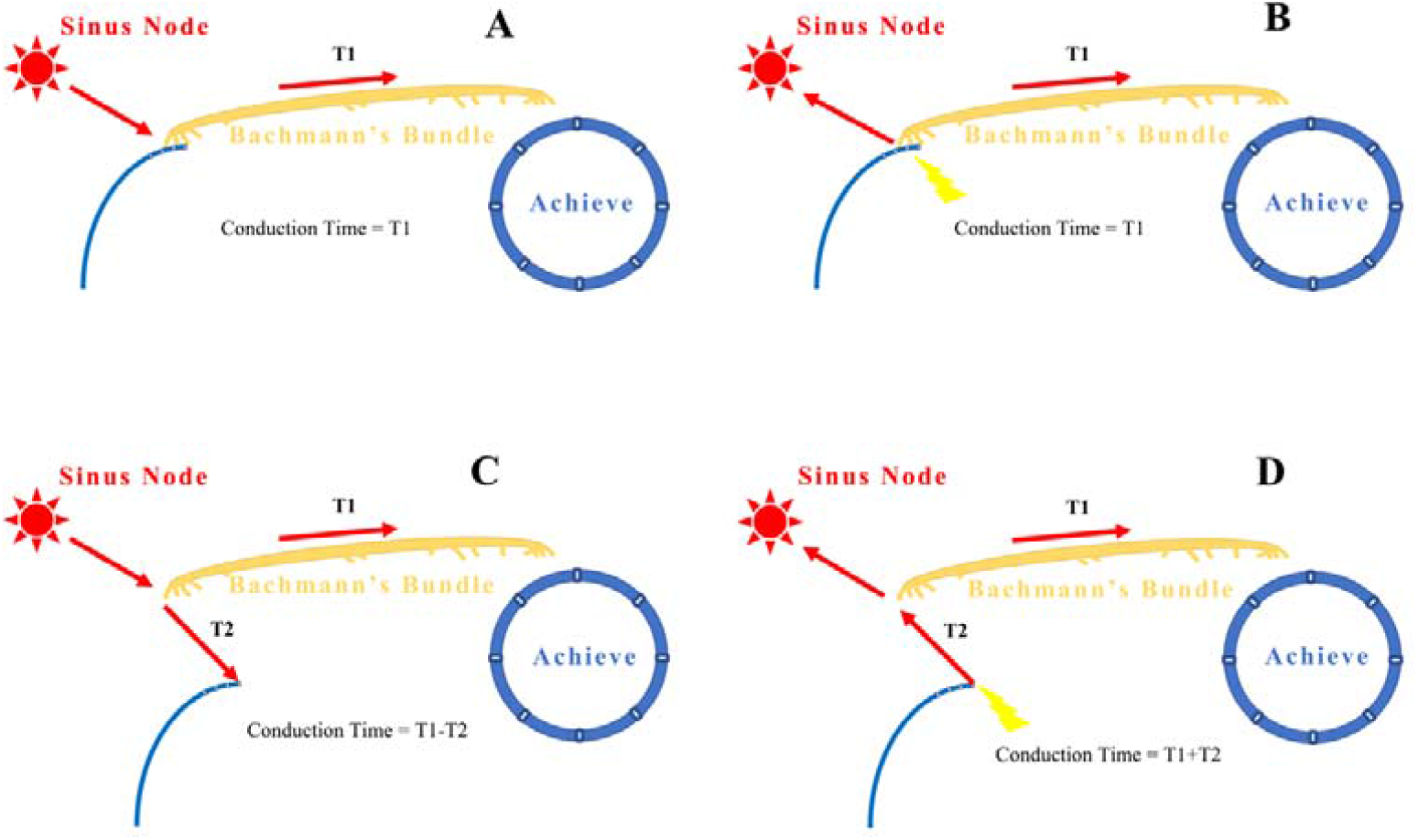
Schematic of ideal pacing position selection. A: In sinus rhythm, Conduction Time = T1 when pacing catheter located on Bachmann’ bundle; B: In pacing rhythm, Conduction Time = T1 when pacing catheter located on Bachmann’ bundle. Conduction Time in sinus rhythm = Conduction Time in pacing rhythm; C: In sinus rhythm, Conduction Time = T1-T2 when pacing catheter located away from Bachmann’ bundle; D: In pacing rhythm, Conduction Time = T1+T2 when pacing catheter located away from Bachmann’ bundle. Conduction Time in sinus rhythm < Conduction Time in pacing rhythm

### 2.5 Follow-up and study end points

Follow-up visits with 24-hour Holter were performed at 3, 6, and 12 months after ablation. Random ECGs were also collected by portable device (the BigThumb™ ECG monitor, Shanghai Yueguang Medical Technologies Inc., Shanghai, China) as we described in another study^13^. AF recurrence is defined as ECG collected by Holter or portable device tracing of more than 30 seconds atrial fibrillation/ atrial flutter/ atrial tachycardia. Primary effectiveness was freedom from AF or atrial tachycardia absent class I/III antiarrhythmic drugs.

### 2.6 Statistical analysis

Continuous variables are expressed as mean ± standard deviation, and categorical variables are expressed as a number and percentage. Continuous variables were compared using the two-tailed Student’s t-test, and categorical variables were compared using the χ 2 test or Fischer’s exact test. In order to examine the difference between groups on AF recurrence, Kaplan-Meier survival curve and log-rank analysis were performed. For Logistic regression investigating factors affecting the success rate of RL block, univariable regression was performed first. Variables with a P-value of < 0.2 were then included in the multivariable model. A level of significance was set at < 0.05 for all reported P-values, and confidence intervals were calculated at the 95% level. All statistics were then calculated using SPSS (version 24, IBM, Armonk, NY, USA).

## 3 Results

### 3.1 Baseline characteristics and ablation parameters

There was no statistical difference in baseline characteristics and pulmonary veins ablation parameters between the PVI group and the PVI+RL group, except for minimum temperature and time to isolation in right superior pulmonary vein (Table 1 and Table 2). In the PVI+RL group, 6.07 ± 2.03 times of freezing sequence were applied for RL block. There was one case of transient phrenic nerve injury during ablation of right superior pulmonary vein in PVI+RL group and the conduction of phrenic nerve gradually returned to normal after cessation of ablation. There was no other serious complication in the two groups.

**Table 1.**
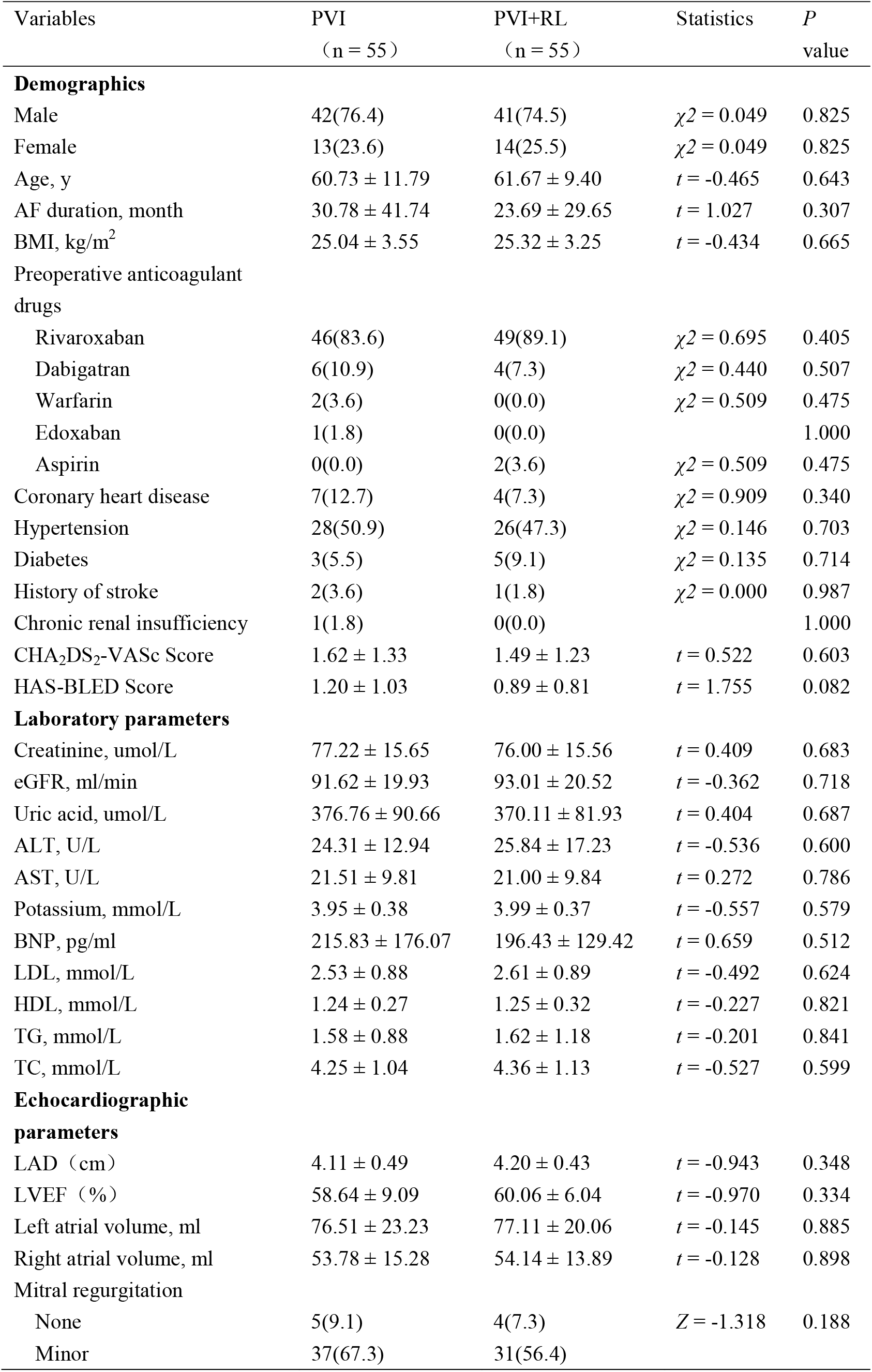
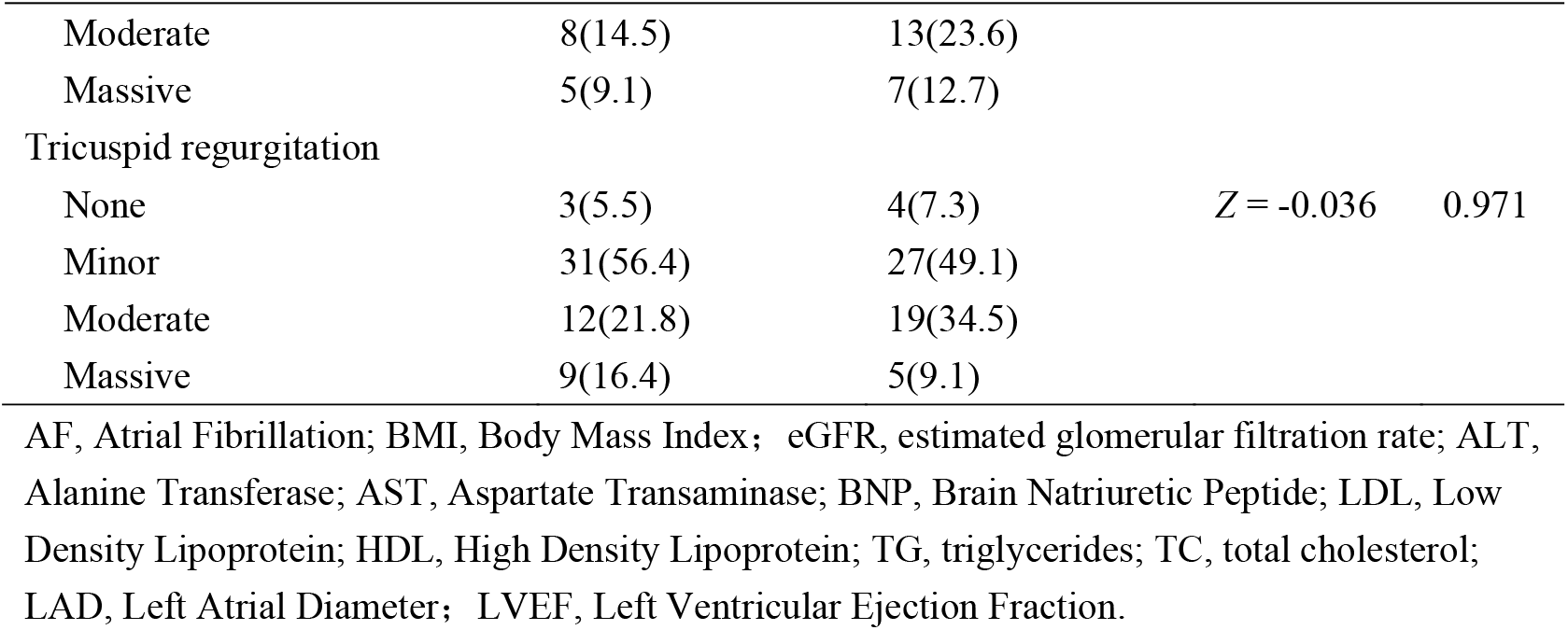
Baseline Characteristics.

**Table 2.** Procedural characteristics.

| Variables | PVI<br>(n = 55) | PVI+RL<br>(n = 55) | Statistics | *P*<br>value |
| --- | --- | --- | --- | --- |
| Procedure time, min | 105.82 ± 25.14 | 145.56 ± 37.37 | *t* = -6.544 | 0.000 |
| Exposure time, min | 12.29 ± 5.98 | 17.69 ± 8.21 | *t* = -3.943 | 0.000 |
| Radiation dose, mGy | 359.64 ± 275.81 | 471.98 ± 389.50 | *t* = -1.746 | 0.084 |
| Freezing times (total) | 7.05 ± 1.99 | 12.33 ± 2.78 | *t* = -11.440 | 0.000 |
| Freezing time, min (total) | 16.62 ± 3.33 | 27.32 ± 5.53 | *t* = -12.278 | 0.000 |
| LSPV |  |  |  |  |
| Freezing times | 2.27 ± 1.03 | 1.98 ± 1.17 | *t* = 1.339 | 0.183 |
| Time to 0°C, s | 12.05 ± 2.20 | 11.94 ± 1.75 | *t* = -0.032 | 0.974 |
| Minimum temperature, °C | -43.67 ± 9.08 | -45.30 ± 7.98 | *t* = 1.355 | 0.178 |
| TTI, s | 44.63 ± 18.22 | 51.58 ± 31.09 | *t* = 1.346 | 0.183 |
| TTI temperature, °C | -38.46 ± 5.63 | -37.30 ± 7.94 | *t* = -0.545 | 0.588 |
| LIPV |  |  |  |  |
| Freezing times | 1.46 ± 0.97 | 1.25 ± 0.52 | *t* = 1.352 | 0.181 |
| Time to 0°C, s | 10.96 ± 1.53 | 10.97 ± 1.83 | *t* = -0.032 | 0.974 |
| Minimum temperature, °C | -42.40 ± 7.35 | -44.05 ± 6.53 | *t* = 1.355 | 0.178 |
| TTI, s | 35.80 ± 23.21 | 27.97 ± 22.92 | *t* = 1.346 | 0.183 |
| TTI temperature, °C | -29.36 ± 8.54 | -28.06 ± 9.70 | *t* = -0.545 | 0.588 |
| RIPV |  |  |  |  |
| Freezing times | 1.88 ± 0.98 | 1.70 ± 0.87 | *t* = 1.031 | 0.305 |
| Time to 0°C, s | 10.21 ± 5.41 | 10.37 ± 1.59 | *t* = -0.257 | 0.797 |
| Minimum temperature, °C | -45.44 ± 7.66 | -44.85 ± 7.62 | *t* = -0.525 | 0.600 |
| TTI, s | 35.15 ± 19.55 | 36.69 ± 17.27 | *t* = -0.301 | 0.765 |
| TTI temperature, °C | -30.16 ± 14.16 | -33.81 ± 9.05 | *t* = 1.092 | 0.281 |
| RSPV |  |  |  |  |
| Freezing times | 2.24 ± 1.44 | 1.77 ± 0.90 | *t* = 1.992 | 0.050 |
| Time to 0°C, s | 10.35 ± 1.35 | 9.55 ± 5.41 | *t* = 1.505 | 0.134 |
| Minimum temperature, °C | -45.24 ± 10.11 | -48.14 ± 7.63 | *t* = 2.313 | 0.022 |
| TTI, s | 34.12 ± 16.08 | 24.39 ± 14.93 | *t* = 2.366 | 0.022 |
| TTI temperature, °C | -32.68 ± 8.24 | -28.35 ± 10.90 | *t* = -1.640 | 0.107 |
| RMPV |  |  |  |  |
| Freezing times | 2.40 ± 2.61 | 1.67 ± 1.16 | *t* = 0.450 | 0.668 |
| Time to 0°C, s | 11.42 ± 1.83 | 9.80 ± 0.84 | *t* = 1.867 | 0.082 |
| Minimum temperature, °C | -39.80 ± 6.26 | -47.40 ± 5.77 | *t* = 1.996 | 0.081 |
| RL |  |  |  |  |
| Freezing times |  | 6.07 ± 2.03 |  |  |
| Time to 0°C, s |  | 10.60 ± 1.85 |  |  |
| Minimum temperature, °C |  | -39.05 ± 5.07 |  |  |

LSPV, Left Superior Pulmonary Vein; LIPV, Left Inferior Pulmonary Vein; RIPV, Right Inferior Pulmonary Vein; RSPV, Right Superior Pulmonary Vein; RMPV, Right Middle Pulmonary Vein; RL, Roof Line; TTI, Time To Isolation.

Three patients in PVI group and one patient in PVI+RL group underwent atrial flutters during or after CBA for PVI which were terminated by radiofrequency ablation for roof line, cavotricuspid isthmus line or anterior wall line. Moreover, AF terminated during PVI for one patient in PVI+RL group, then additional RF ablation was abandoned.

### 3.2 Primary outcome

There was no significant difference in atrial arrhythmia-free survival rate between PVI group and PVI+RL group (63.5% vs 76.2%, P = 0.296, HR = 0.707, 95%CI = [0.366, 1.366]) after 532.7 ± 171.0 days of follow-up (Figure 4). However, atrial arrhythmia-free survival rate in patients with blocked RL was extremely improved compared with those unblocked (84.0% vs 45.5%, P = 0.023, HR 0.332, 95%CI = [0.120, 0.916]) as shown in Figure 5.

**Figure 4.**
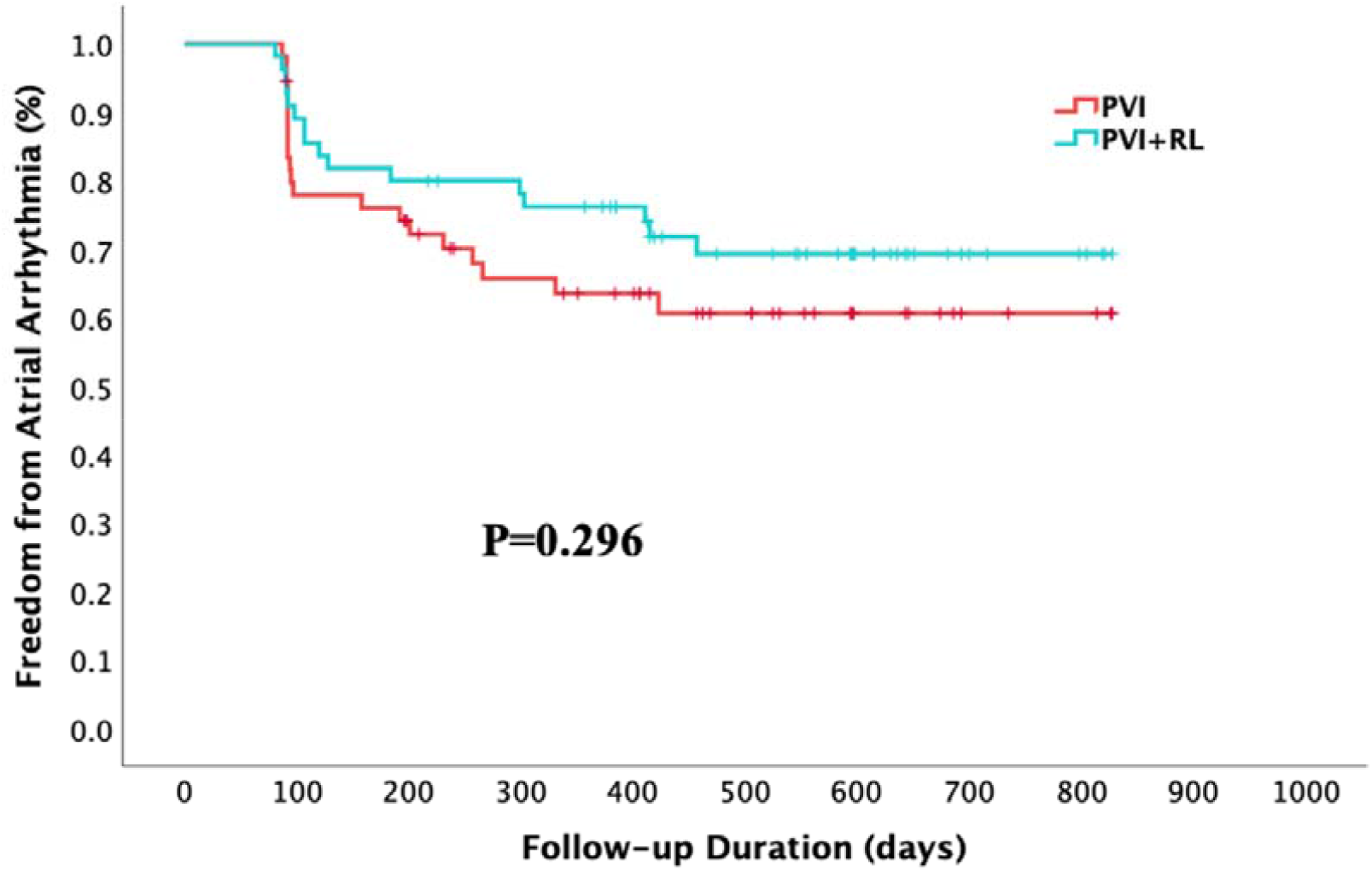
The Kaplan-Meier survival curve for the primary endpoint in PVI and PVI+RL group.

**Figure 5.**
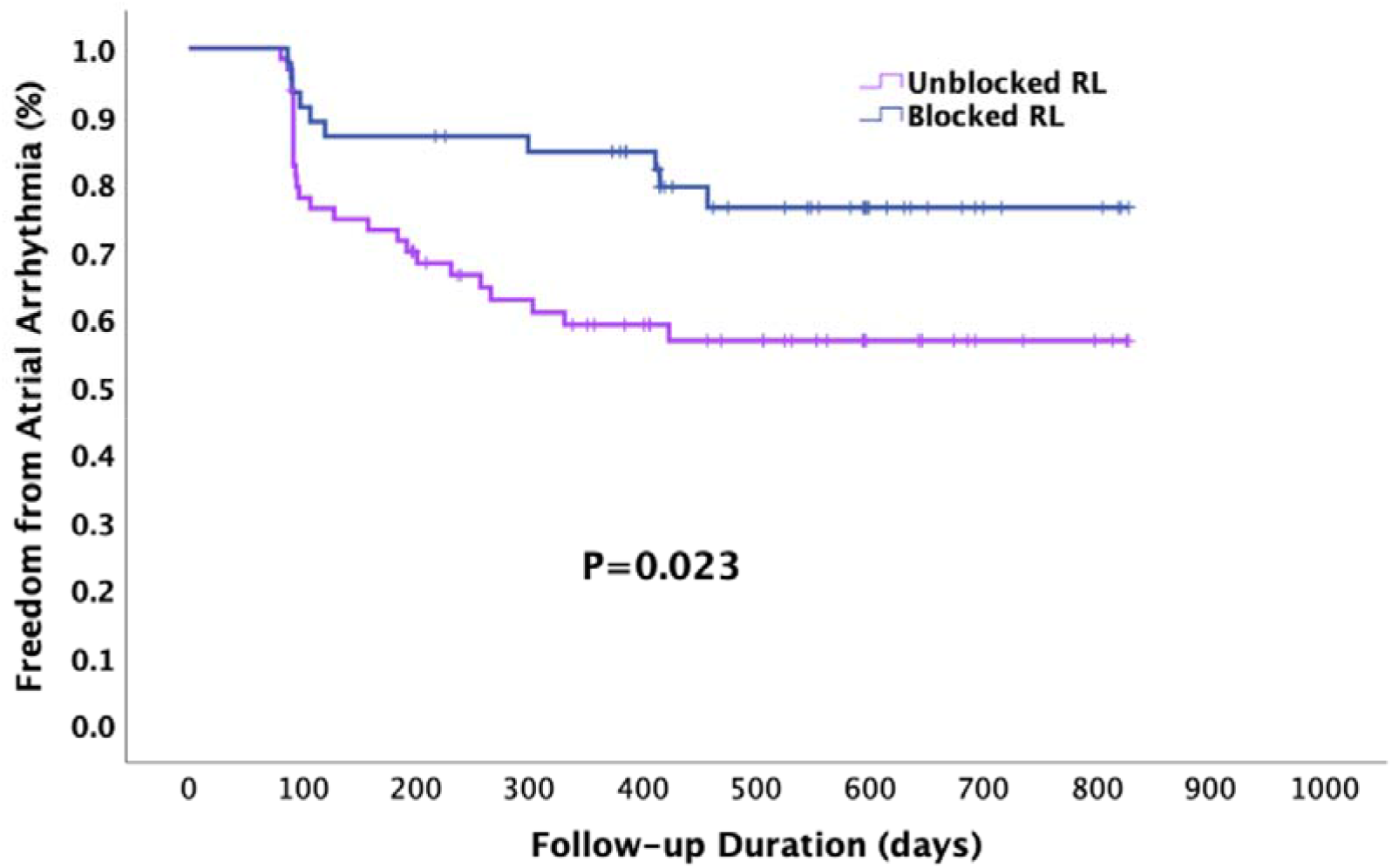
The Kaplan-Meier survival curve for the primary endpoint in patients with blocked and unblocked roof line.

### 3.3 RL block assessment

Upper right atrial septum pacing and activation mapping of posterior wall of left atrium verified all the block and revealed all the connection position of RL. Activation mapping of posterior wall of left atrium in sinus rhythm also verified the block of most of the RLs (53/55, 96.4%). However, the accuracy rate of voltage mapping was only 87.3% (48/55).

There were 38/55 patients were achieved RL block after initial attempt of RL CBA with abandon of RL ablation in one patient. The connection positions of the RL were revealed in the middle of RL in 13 patients and additional ablation achieved RL block in 6 patients. RL block could not be achieved even with additional ablation in the remaining 6 patient. There were still two patients with connection positions in right part of RL and one patient in left part which could not be blocked after additional ablation. The success rate of RL block in PVI+RL group was 80% (44/55).

### 3.4 Factors affecting RL block

Univariate Logistic regression analysis showed that the factors affecting RL complete block were RL angle (HR 1.04, 95%CI 1.00-1.08, P = 0.056) and RL shape (HR 6.81, 95%CI 1.63-28.45, P = 0.009). Multivariate Logistic regression analysis showed that RL shape was the only factor affecting RL complete block (HR 4.79, 95%CI 1.02-22.58, P = 0.048) as shown in Table 3.

**Table 3.**
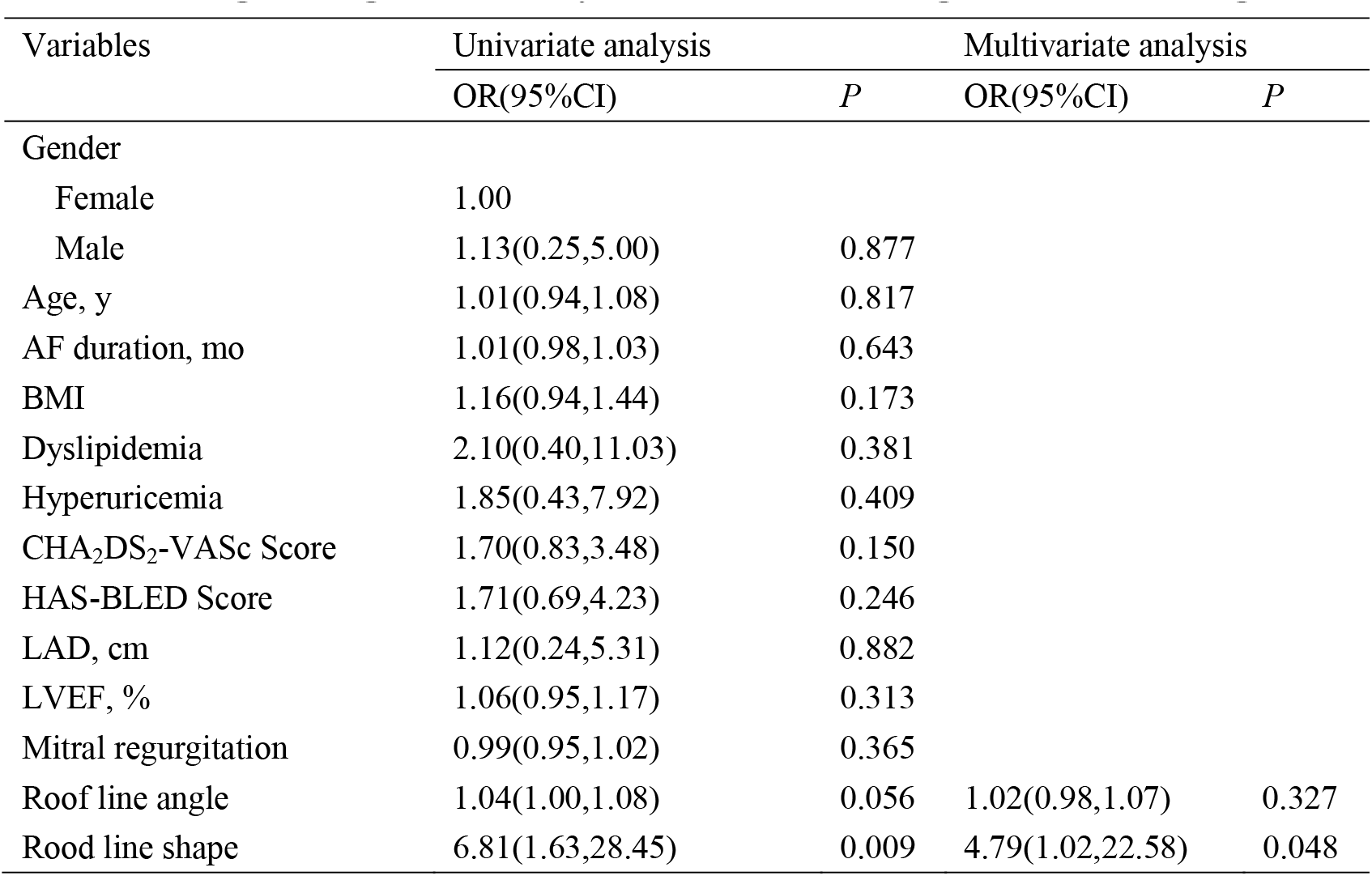
Logistic regression analysis of factors affecting roof line blocking rate.

RL shape was categorized in ‘Regular shape’ and ‘Irregular shape’. ‘Regular shape’ was defined as flat RL with left part slightly higher which was the most common shape of RL. ‘Irregular shape’ included RL with shape of ‘V’, ‘W’, ‘Ω’, ‘L’, ‘-’, and so on (Figure 6). The rate of complete block of RL was significantly high in ‘Regular shape’ group (35/39, 89.7%) than ‘Irregular shape’ group (9/16, 56.3%, P = 0.014).

**Figure 6.**
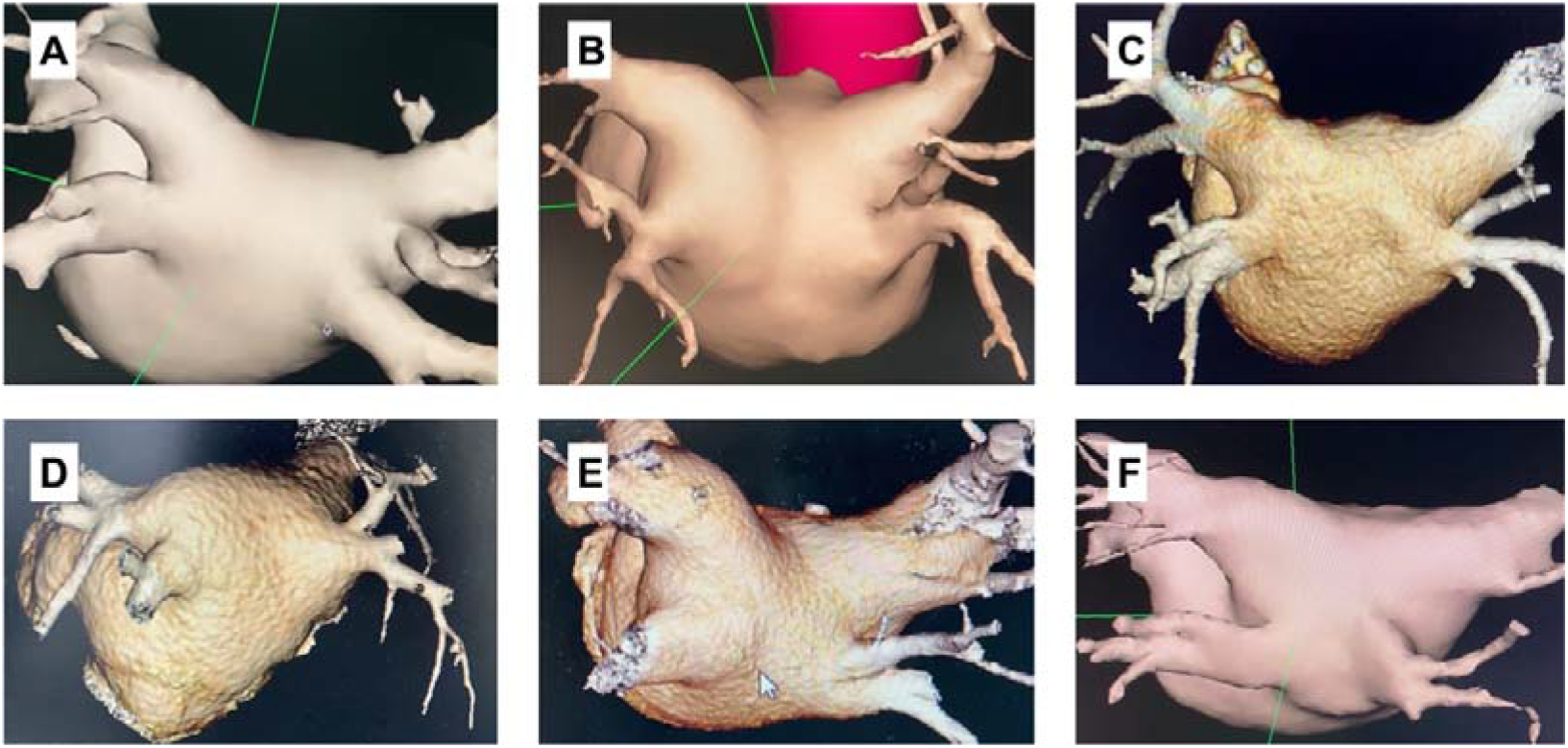
The categorization of roof line shape. A: ‘Regular shape’, flat RL with left part slightly higher; B: ‘V shape’, the middle of RL was the lowest with the RL like a ‘V’; C: ‘W shape’, the RL was not flat like a ‘W’; D: ‘Ω shape’, the middle of RL was the highest with the RL like a ‘Ω’; E: ‘L shape’, there was a corner in RL close to left or right superior pulmonary vein.; F: ‘-shape’, the RL was flat and horizontal with almost the same height in left and right part of RL

## Discussion

To our knowledge, this is the first randomized controlled study to prospectively demonstrate the efficacy of a novel CBA strategy for block of roof line and reduction of atrial fibrillation recurrence. We found that blocked roof line ablation was associated with a significant reduction in risk of atrial fibrillation recurrence after CBA. Patients with ‘Regular’ shape of roof line may benefit more from roof line ablation.

Accumulating evidence have demonstrated that RL ablation in addition to PVI reduce AF recurrence significantly^6, 7, 14^. The area modified by ablation was extremely larger in cryoballoon than radiofrequency ablation^15, 16^. It implied that RL block achieved by CBA may further reduce AF recurrence compared to radiofrequency^17-19^.

The CARFI-PerAF study found that blocked RL ablation could improve success rate. However, 54.5% of patients with unblocked RL suffered from AF recurrence after CBA. So, it is extremely important to improve the block rate of RL ablation, which was reported to be 88.2%-91.9% with a significantly higher AF-free rate after follow-up^9, 20^. However, it is challenging to achieve and verify RL complete block using cryoballoon because the contact between the cryoballoon and RL was not intuitively observed.

‘Quarter balloon ablation technique’ and ‘roof distortion technique’ were used to improve quality of RL ablation in this study which may result in more effective contact and extensive lesion by the cryoballoon. Moreover, conduction block of RL was confirmed by both voltage mapping and upper right atrial septum pacing. It seems that the conduction block rate may be overestimated by the voltage mapping alone. With effort, the block rate of RL in the CARFI-PerAF was only 80%. Logistic regression analysis showed that RL shape was the only factor affecting RL complete block. The block rate was extremely high in patients with ‘Regular shape’ RL. In contrast to that, ‘Irregular shape’ predicted a lower conduction block rate with a RL shape of ‘V’, ‘W’, ‘Ω’, ‘L’, ‘-’, and so on. This suggested that patients with ‘Regular shape’ RL may benefit more from RL cryoballoon ablation.

### Limitations

This study has limitations. First, this study is a single center clinical trial with Chinese patients as object. ‘Quarter balloon ablation technique’ and ‘roof distortion technique’ needs promotion to become widespread which was not generally used in other centers. Thus, a multi-center study should be needed to validate the study findings. The second is small sample size that may lead to the nonsignificance in AF recurrence between the two groups. The sample size was calculated according to success rate reported by previous literatures that made a difference with the CARFI-PerAF study. Further studies are necessary with large sample size to confirm the reduction in AF recurrence with RL cryoballoon ablation.

## Data Availability

Due to the nature of this research, participants of this study did not agree for their data to be shared publicly, so supporting data is not available.

